# Longitudinal Associations of Cigarette Smoking, Smoking Intensity, and Smoking Trajectories With Emergency Room Utilization Among U.S. Adults

**DOI:** 10.64898/2026.10.31.26366988

**Authors:** Wanru Guo, Xinyu Yan, Curtis Tatsuoka

## Abstract

**Background:** Tobacco smoking is a leading cause of morbidity and mortality in the United States, yet its longitudinal association with emergency room (ER) utilization remains understudied. We examined whether cigarette smoking, smoking intensity, and smoking trajectories were associated with subsequent ER use.

**Methods:** We analyzed adult respondents from Waves 1–6 of the Population Assessment of Tobacco and Health Study. Wave 1–3 smoking status, intensity, and cigarettes/day were examined in relation to next-wave ER outcomes. Logistic generalized estimating equation models were used for any ER visit and frequent ER use (*≥*2 visits), and quasi-Poisson models were used for ER visit count. Trajectory analyses examined Wave 1–5 smoking patterns and Wave 6 ER outcomes, with Wave 1–4 to Wave 5 as sensitivity analysis. Marginal structural models and propensity score matching were used to strengthen causal interpretation. Models adjusted for demographics, insurance, self-rated health, and baseline ER use. Survey weights with balanced repeated replication and Fay’s adjustment were used.

**Results:** In adjusted GEE models, current smoking was associated with higher subsequent ER utilization, including any ER visit (OR 1.128, 95% CI [1.068–1.191]), frequent ER use (OR 1.275, 95% CI [1.185–1.372]), and ER visit count (RR 1.264, 95% CI [1.160–1.377]; all p<0.001). Smoking intensity showed a dose-response pattern: smoking <20 cigarettes/day was associated with any ER visit (OR 1.130, 95% CI [1.060–1.204]), frequent ER use (OR 1.231, 95% CI [1.124–1.347]), and ER visit count (RR 1.262, 95% CI [1.110–1.435]), while smoking *≥*20 cigarettes/day showed stronger associations with any ER visit (OR 1.231, 95% CI [1.127–1.345]), frequent ER use (OR 1.447, 95% CI [1.293–1.618]), and ER visit count (RR 1.368, 95% CI [1.245–1.504]; all p<0.001). Each additional cigarette/day was also associated with higher ER utilization (OR/RR range, 1.007– 1.011; all p<0.001). Findings were consistent in marginal structural models and propensity score matched analyses. In trajectory analyses, new/intermittent and persistent smokers had significantly higher ER utilization, while former/intermittent smokers and sustained quitters did not differ from non-smokers, suggesting that this harmful association was eliminated by smoking cessation.

**Conclusions:** Smoking, heavier intensity, and persistent or emerging smoking patterns were associated with higher subsequent ER utilization, while smoking cessation reduces ER risk to the same level as non-smokers. This supports smoking prevention and cessation as strategies to reduce acute healthcare burden.

**IMPLICATIONS:** Smoking should be considered not only a risk factor for long-term chronic disease, but also a predictor of acute healthcare utilization. The consistency of findings across longitudinal, marginal structural, and propensity score matched analyses supports a potential causal relationship between smoking exposure and subsequent emergency room use. These results strengthen the rationale for integrating smoking cessation counseling, nicotine replacement therapy, quitline referral, and preventive care linkage into emergency and urgent care settings, alongside broader tobacco-control policies to reduce acute healthcare burden.

## INTRODUCTION

Tobacco use remains a major public health burden in the United States and globally. In the United States, cigarette smoking and secondhand smoke exposure cause more than 480,000 deaths each year, nearly one in five deaths, and cost more than $600 billion in 2018, including more than $240 billion in health care spending (CDC, 2024). Although adult cigarette smoking has declined, with 9.9% of U.S. adults reporting current cigarette smoking in 2024 compared with 11.6% in 2022, tobacco-related morbidity remains substantial (CDC, 2024; NCHS, 2026). Globally, tobacco causes more than 7 million deaths each year, including more than 1.6 million deaths among nonsmokers exposed to secondhand smoke (WHO, 2026).

Emergency room (ER) care represents an important but understudied setting for understanding the acute healthcare burden of smoking. In 2022, the United States had approximately 155.4 million emergency department visits, or 47.3 visits per 100 people (CDC/NCHS, 2024). National ED visit costs totaled $76.3 billion across 144.8 million visits in 2017, and claims-based data show continued increases in ER spending and prices (Moore & Liang, 2020; Health Care Cost Institute, 2023). Smokers may be overrepresented in emergency care; although current smoking prevalence among U.S. adults is below 14%, smokers may account for approximately 30–40% of ED patients (Yale Medicine, n.d.), yet the longitudinal relationship between smoking and ER utilization remains insufficiently characterized.

Prior studies have linked smoking with increased ED utilization, particularly among patients with asthma, diabetes, chronic obstructive pulmonary disease, and respiratory disease (Barmparessou et al., 2020; Selya et al., 2020; Madani & Carpenter, 2023). Cross-sectional ED studies have also shown that tobacco use is common among ED patients and that the ED may be an important setting for identifying smokers and delivering cessation interventions (Smith, 2011). More recent evidence further supports this clinical relevance: smoking has been associated with frequent treat– and-release ED visits across health-complexity groups (Castner et al., 2015), and an ED-based cessation trial improved 6-month biochemically verified abstinence compared with usual care (Pope et al., 2024). However, much of the existing literature remains disease-specific, geographically limited, retrospective, or cross-sectional, limiting generalizability and causal interpretation.

To address these gaps, this study examined the longitudinal association between cigarette smoking and subsequent ER utilization among U.S. adults, including any ER visit, frequent ER use, and ER visit count. We also evaluated whether smoking intensity and smoking behavior trajectories, including persistent smoking, new/intermittent smoking, former/intermittent smoking, sustained quitting, and relapse, were associated with later ER outcomes. Using data from the nationally representative Population Assessment of Tobacco and Health (PATH) Study, this analysis aimed to clarify how smoking status, smoking intensity, and smoking patterns over time relate to acute healthcare utilization.

## METHODS

### Study population

The PATH Study is a nationally representative longitudinal cohort study of U.S. adults and youth. The study used a four-stage, stratified, address-based probability sampling design and oversampled adult tobacco users, young adults aged 18–24 years, and Black adults. Full-sample and replicate weights were created to account for the complex sampling design, oversampling, and nonresponse (Hyland et al., 2017; Opsomer et al., 2024).

The primary longitudinal analysis used smoking exposures from Waves 1–3 to predict ER outcomes at the subsequent wave, corresponding to Wave 1 to Wave 2, Wave 2 to Wave 3, and Wave 3 to Wave 4. Secondary trajectory analyses examined smoking behavior patterns from Waves 1–5 in relation to Wave 6 ER outcomes, with sensitivity analyses using Waves 1–4 smoking patterns to predict Wave 5 ER outcomes during the pre-pandemic period.

### Predictors and outcomes

Smoking exposure was defined using PATH-derived current established cigarette smoking status and average cigarettes smoked per day. Current established cigarette smoking was based on the PATH-derived variable R0#R_A_CUR_ESTD_CIGS. Smoking intensity was categorized among current established smokers as <20 cigarettes/day or *≥*20 cigarettes/day using the PATH-derived average cigarettes-per-day variable; non-smokers served as the reference group. ER outcomes were based on self-reported emergency room or urgent care use in the past 12 months, including any ER visit (R0#_AX0302), frequent ER use defined as *≥*2 visits, and total ER visit count (R0#_AX0155).

For the primary longitudinal analysis, smoking exposures were measured at Waves 1–3 and linked to ER outcomes at the subsequent waves. For trajectory analyses, smoking patterns were defined across Waves 1–5, with Waves 1–4 used in sensitivity analyses. Trajectory categories included persistent non-smokers, persistent smokers, former/intermittent smokers, new/intermittent smokers, sustained quitters, and relapsed/intermittent smokers. Persistent non-smokers were non-smokers at all included waves and served as the reference group. Persistent smokers were smokers at all included waves. Former/intermittent smokers were smokers at Wave 1 but were not smokers at all subsequent waves. New/intermittent smokers were non-smokers at Wave 1 but reported smoking at one or more later waves. Sustained quitters were Wave 1 smokers who remained non-smokers at all subsequent included waves, and relapsed/intermittent smokers were Wave 1 smokers with mixed smoking status across later waves.

### Baseline measures

Baseline covariates were selected to capture demographic, socioeconomic, healthcare access, and health-status factors previously associated with tobacco use and ER utilization. These included age group, sex, race/ethnicity, education, income, insurance status, self-rated physical health, and baseline ER visit count. Reference groups were age 18–24 years, male sex, White race/ethnicity, less than high school education, income <$10,000, no insurance, and excellent self-rated physical health. Education categories included GED, high school graduate, some college/associate degree, bachelor’s degree, and advanced degree. Income categories included $10,000–$24,999, $25,000–$49,999, $50,000–$99,999, and *≥*$100,000. Insurance categories included private insurance, Medicaid, Medicare and other insurance.

### Statistical analysis

#### Primary longitudinal association analysis

For the primary longitudinal analysis, current established cigarette smoking status, smoking intensity category, and cigarettes smoked per day were examined as time-varying exposures across Waves 1–3, with ER outcomes assessed at the subsequent wave (2-4). Covariates adjusted included prior ER utilization, self-rated health, and sociodemographic factors, including age, sex, race/ethnicity, education, income, and insurance status. Generalized estimating equation models were used to account for repeated observations within participants (Liang & Zeger, 1986). Logistic GEE models were used for binary outcomes, including any ER visit and frequent ER use defined as two or more ER visits, and negative binomial GEE models were used for ER visit count to account for the zero-inflated nature of the visits (Table 1).

**Table 1:** Wave 1 baseline characteristics by current established smoking status.

| Characteristic | Overall | Current established smoker | Non-smoker |
|---|---|---|---|
| **Age, years: 18 to 24** | 9,098 (28.2) | 2,477 (21.7) | 6,621 (31.8) |
| 25 to 34 | 6,332 (19.6) | 2,589 (22.7) | 3,743 (18.0) |
| 35 to 44 | 4,917 (15.3) | 2,048 (18.0) | 2,869 (13.8) |
| 45 to 54 | 4,828 (15.0) | 2,051 (18.0) | 2,777 (13.3) |
| 55 to 64 | 3,958 (12.3) | 1,515 (13.3) | 2,443 (11.7) |
| 65 to 74 | 2,111 (6.5) | 578 (5.1) | 1,533 (7.4) |
| 75 or older | 989 (3.1) | 143 (1.3) | 846 (4.1) |
| **Sex: Male** | 16,276 (50.5) | 5,986 (52.5) | 10,290 (49.4) |
| Female | 15,963 (49.5) | 5,416 (47.5) | 10,547 (50.6) |
| **Race/ethnicity: White alone** | 23,832 (73.9) | 8,700 (76.3) | 15,132 (72.6) |
| Black alone | 5,031 (15.6) | 1,610 (14.1) | 3,421 (16.4) |
| Other | 3,376 (10.5) | 1,092 (9.6) | 2,284 (11.0) |
| **Education: Less than high school** | 4,210 (13.1) | 1,896 (16.7) | 2,314 (11.2) |
| GED | 2,209 (6.9) | 1,285 (11.3) | 924 (4.5) |
| High school graduate | 7,526 (23.5) | 2,876 (25.4) | 4,650 (22.5) |
| Some college or associate degree | 11,286 (35.2) | 4,065 (35.9) | 7,221 (34.9) |
| Bachelor’s degree | 4,494 (14.0) | 896 (7.9) | 3,598 (17.4) |
| Advanced degree | 2,309 (7.2) | 305 (2.7) | 2,004 (9.7) |
| **Household income: Less than \$10,000** | 5,649 (19.4) | 2,411 (23.1) | 3,238 (17.4) |
| \$10,000 to \$24,999 | 6,750 (23.2) | 2,979 (28.5) | 3,771 (20.2) |
| \$25,000 to \$49,999 | 6,657 (22.9) | 2,590 (24.8) | 4,067 (21.8) |
| \$50,000 to \$99,999 | 6,133 (21.1) | 1,784 (17.1) | 4,349 (23.3) |
| \$100,000 or more | 3,912 (13.4) | 690 (6.6) | 3,222 (17.3) |
| **Insurance status: No insurance** | 6,137 (19.3) | 2,968 (26.3) | 3,169 (15.4) |
| Some private insurance | 19,354 (60.8) | 5,349 (47.4) | 14,005 (68.1) |
| No private insurance or Medicare, some Medicaid | 3,158 (9.9) | 1,613 (14.3) | 1,545 (7.5) |
| No private insurance, some Medicare | 2,088 (6.6) | 951 (8.4) | 1,137 (5.5) |
| Other insurance only | 1,104 (3.5) | 402 (3.6) | 702 (3.4) |
| **Self-rated physical health: Excellent** | 4,883 (15.2) | 969 (8.5) | 3,914 (18.8) |
| Very good | 10,574 (32.8) | 2,922 (25.7) | 7,652 (36.8) |
| Good | 11,026 (34.2) | 4,539 (39.9) | 6,487 (31.2) |
| Fair | 4,634 (14.4) | 2,326 (20.4) | 2,308 (11.1) |
| Poor | 1,078 (3.3) | 629 (5.5) | 449 (2.2) |
| **ER/urgent care visit at Wave 1: No** | 23,176 (72.1) | 7,576 (66.6) | 15,600 (75.0) |
| Yes | 8,989 (27.9) | 3,794 (33.4) | 5,195 (25.0) |
| **Frequent ER/urgent care use at Wave 1 (≥2 visits): No** | 28,217 (87.8) | 9,531 (83.9) | 18,686 (89.9) |
| Yes | 3,924 (12.2) | 1,826 (16.1) | 2,098 (10.1) |
| **Number of ER/urgent care visits at Wave 1, mean (SD)** | 0.6 (1.9) | 0.7 (2.2) | 0.5 (1.7) |
| Median [IQR] | 0.0 [0.0, 1.0] | 0.0 [0.0, 1.0] | 0.0 [0.0, 0.0] |

Values are n (%) unless otherwise indicated.

#### Causal analysis using marginal structural models

As a causal sensitivity analysis, marginal structural models were fit using inverse probability weighting to account for time-varying smoking exposure and time-varying confounding (Robins et al., 2000). For the next-wave smoking analysis, stabilized inverse probability weights were estimated from models for smoking exposure conditional on prior smoking status, prior ER utilization, self-rated health, and sociodemographic factors, including age, sex, race/ethnicity, education, income, and insurance status. Weighted generalized estimating equation models were then used to estimate the marginal association between smoking exposure at Waves 1–3 and ER outcomes at the subsequent wave. Outcomes included any ER visit, frequent ER use defined as two or more ER visits, and ER visit count.

#### Propensity score matching

As a design-based sensitivity analysis, propensity score matching was used to compare current established smokers with non-smokers within each next-wave interval (Rosenbaum & Rubin, 1983). Matching was performed separately for Wave 1 smoking predicting Wave 2 ER outcomes, Wave 2 smoking predicting Wave 3 ER outcomes, and Wave 3 smoking predicting Wave 4 ER outcomes. Propensity scores were estimated using age, sex, race/ethnicity, education, income, insurance status, self-rated physical health, and prior ER visit count. Matched samples were then analyzed for three subsequent-wave outcomes: any ER visit, frequent ER use defined as two or more ER visits, and ER visit count.

#### Smoking pattern analysis

For the longer-term trajectory analysis, smoking exposure patterns from Waves 1–5 were used to predict ER outcomes at Wave 6 for both the former/intermittent smoking analysis and the sustained quitter analysis. As a secondary sensitivity analysis, smoking patterns from Waves 1–4 were used to predict Wave 5 ER outcomes for sustained quitter analysis. Multivariable logistic regression models were used for binary outcomes and Poisson regression models were used for ER visit count. Models were adjusted for Wave 1 ER visit count, Wave 1 self-rated physical health, age, sex, race/ethnicity, education, income, and insurance status, with persistent non-smokers serving as the reference group. As a causal sensitivity analysis, marginal structural models were fit using stabilized inverse probability weights. The trajectory weight model included the same baseline covariates, and weighted logistic and Poisson models were used to estimate Wave 6 ER outcomes.

All analyses in this study were done using R version 4.6.1 (R Development Core Team, Vienna, Austria). Weighted analyses incorporated PATH full-sample and replicate weights to account for the complex survey design. Point estimates were obtained using the full-sample weight, and variances were estimated from 100 replicate-weighted fits using balanced repeated replication with Fay’s adjustment ( = 0.30). Results are presented as odds ratios for binary outcomes and rate ratios for count outcomes, with 95% confidence intervals and two-sided p-values.

## RESULTS

### Demographics

At Wave 1, the mean age was 28.2 years; 28.2% were aged 18–24, 19.6% 25–34, 15.3% 35–44, 15.0% 45–54, 12.3% 55–64, 6.5% 65–74, and 3.1% 75 years or older. The sample was 50.5% male and 49.5% female, and was predominantly White (73.9%), followed by Black (15.6%) and other race/ethnicity groups (10.5%). The most common education category was some college or associate degree (35.2%), followed by high school graduate (23.5%) and bachelor’s degree (14.0%). Most respondents reported excellent, very good, or good self-rated physical health, although fair or poor health was more common among current established smokers.

At baseline, 27.9% reported an ER or urgent care visit in the past 12 months, 12.2% reported frequent ER use, and the mean number of ER/urgent care visits was 0.6. Baseline ER/urgent care use was higher among current established smokers than non-smokers, including any ER/urgent care visit (33.4% vs. 25.0%), frequent ER use (16.1% vs. 10.1%), and mean number of visits (0.7 vs. 0.5) (Table 1).

### Longitudinal association between smoking and ER visits

Because smokers and non-smokers differed in baseline demographics and ER utilization (Table 1), models were adjusted for prior ER use, self-rated health, age, sex, race/ethnicity, education, income, and insurance status. Compared with non-smokers, current established smokers had higher subsequent ER utilization, including any ER visit (OR 1.128, 95% CI [1.068–1.191]), frequent ER use (OR 1.275, 95% CI [1.185–1.372]), and ER visit count (RR 1.264, 95% CI [1.160–1.377]; all p<0.001) (Figure 1D; Supplementary Table 1A–C).

**Figure 1.**
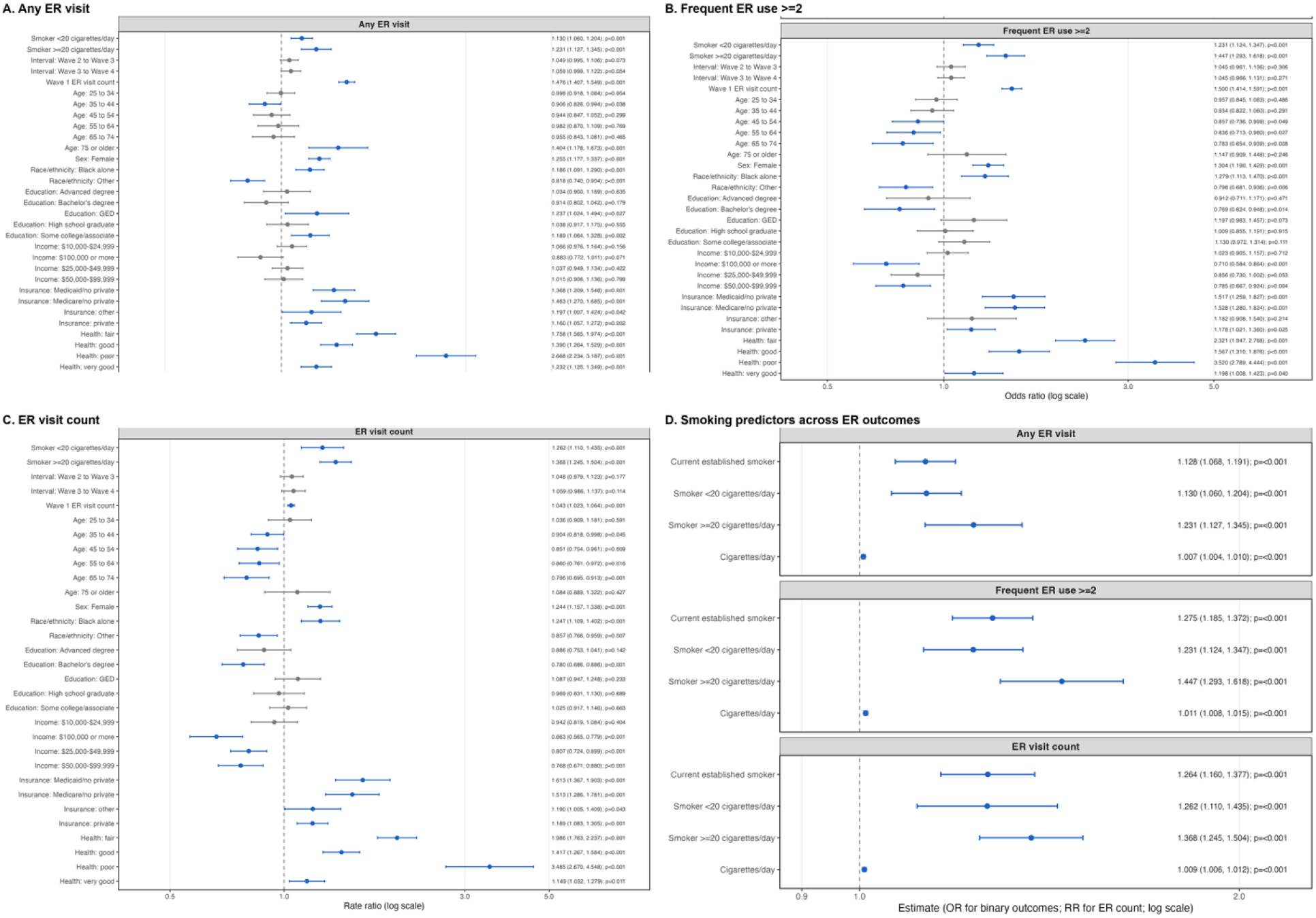
Adjusted associations between smoking and next-wave emergency room utilization. Weighted BRR/Fay estimates are shown for any ER visit, frequent ER use (*≥*2 visits), and ER visit count. Panels A–C show full covariate-adjusted models for smoking intensity; Panel D summarizes smoking status, smoking intensity, and cigarettes/day across outcomes. Full covariate-adjusted models for smoking status and cigarettes/day are shown in Supplementary Figure 1A–C and 1D–F, respectively. Points represent adjusted estimates, horizontal bars represent 95% confidence intervals, and dashed vertical lines indicate the null value of 1.0.

A dose-response pattern was observed for smoking intensity. Compared with non-smokers, smokers consuming *≥*20 cigarettes/day had consistently higher ORs/RRs than those consuming <20 cigarettes/day across all three outcomes. For any ER visit, the OR was 1.231 (95% CI [1.127– 1.345]) for *≥*20 cigarettes/day and 1.130 (95% CI [1.060–1.204]) for <20 cigarettes/day. For frequent ER use, the corresponding ORs were 1.447 (95% CI [1.293–1.618]) and 1.231 (95% CI [1.124–1.347]), and for ER visit count, the RRs were 1.368 (95% CI [1.245–1.504]) and 1.262 (95% CI [1.110–1.435]), respectively; all p<0.001 (Figure 1D–F).

Each additional cigarette/day was also associated with higher ER utilization after covariate adjustment, including any ER visit (OR 1.007, 95% CI [1.004–1.010]), frequent ER use (OR 1.011, 95% CI [1.008–1.015]), and ER visit count (RR 1.009, 95% CI [1.006–1.012]; all p<0.001) (Figure 1D; Supplementary Figure 1D–F). Per additional pack/day, this corresponded to a 15% increase in odds of any ER visit (OR 1.15, 95% CI [1.08–1.22]), a 24% increase in odds of frequent ER use (OR 1.24, 95% CI [1.17–1.35]), and a 20% increase in the rate of ER visits (RR 1.20, 95% CI [1.13–1.27]).

To further evaluate the potential causal association, we used marginal structural models with inverse probability weighting to account for time-varying smoking exposure and time-varying con-founding. Figure 2 presents Fay’s BRR marginal structural model estimates for smoking predictors and subsequent ER outcomes. For any ER visit, current established smoking was associated with higher odds (OR 1.305, 95% CI [1.236–1.377]), as were smoking <20 cigarettes/day (OR 1.317, 95% CI [1.249–1.390]), smoking *≥*20 cigarettes/day (OR 1.486, 95% CI [1.363–1.621]), and each additional cigarette/day (OR 1.015, 95% CI [1.012–1.018]; all p<0.001). For frequent ER use (*≥*2 visits), the corresponding ORs were 1.689, 1.656, 2.026, and 1.024 per cigarette/day, respectively.

**Figure 2.**
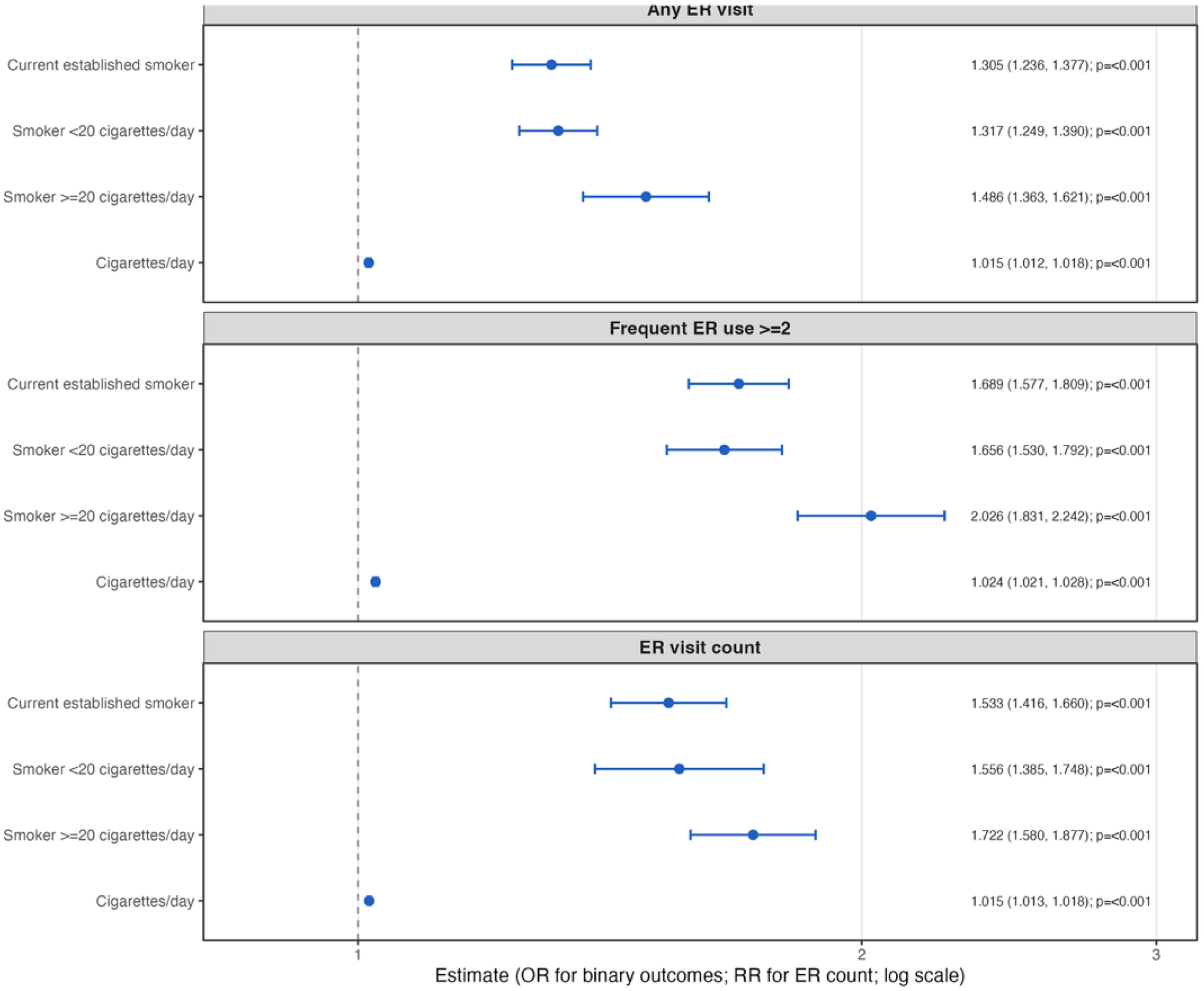
Marginal structural model estimates of smoking exposure and next-wave ER utilization. Estimates account for measured time-varying confounding through stabilized inverse-probability weighting. Error bars indicate 95% confidence intervals calculated using BRR with Fay’s adjustment.

For ER visit count, the corresponding RRs were 1.533, 1.556, 1.722, and 1.015 per cigarette/day, respectively; all associations were significant at p<0.001. These findings support a potential causal relationship between smoking exposure and subsequent ER utilization.

#### Sensitivity analysis

Because several covariates were strongly associated with ER utilization, we conducted additional sensitivity analyses to assess the robustness of the smoking-ER association. Propensity scores included age, sex, race/ethnicity, education, income, insurance status, self-rated health, and prior ER visit count. In the matched analyses, smoking remained associated with higher odds of any ER visit from Wave 1 to Wave 2 (OR 1.183, 95% CI [1.091–1.283], p<0.001) and Wave 2 to Wave 3 (OR 1.170, 95% CI [1.056–1.297], p=0.003). Smoking was also consistently associated with frequent ER use across all intervals, with ORs ranging from 1.178 to 1.360. For ER visit count, associations were significant from Wave 1 to Wave 2 (RR 1.183, 95% CI [1.083–1.291], p<0.001) and Wave 2 to Wave 3 (RR 1.224, 95% CI [1.112–1.347], p<0.001) (Supplementary Figure 2).

### Longer-term smoking trajectories and ER utilization

To evaluate longer-term smoking patterns, we used smoking exposures across Waves 1–5 to define trajectory groups, with persistent non-smokers serving as the reference group. For any ER visit, new/intermittent smokers had higher odds (OR 1.292, 95% CI [1.053–1.585], p=0.014), as did persistent smokers (OR 1.161, 95% CI [1.043–1.292], p=0.006), while former/intermittent smokers were not significantly different from persistent non-smokers (OR 1.038, 95% CI [0.879–1.225], p=0.662). (Figure 3A, summarized in Figure 3D). Similar patterns were observed for frequent ER use (new/intermittent: OR 1.403, 95% CI [1.107–1.779], p=0.005; persistent: OR 1.231, 95% CI [1.040–1.457], p=0.016; former/intermittent: OR 1.172, 95% CI [0.935–1.470], p=0.169) and ER visit count (new/intermittent: RR 1.294, 95% CI [1.083–1.545], p=0.004; persistent: RR 1.254, 95% CI [1.080–1.456], p=0.003; former/intermittent: RR 1.165, 95% CI [0.967–1.404], p=0.108). (Figure 3 B-D). This suggests that individuals who stopped smoking after Wave 1 had ER utilization patterns closer to persistent non-smokers than to continued or newly smoking groups.

**Figure 3.**
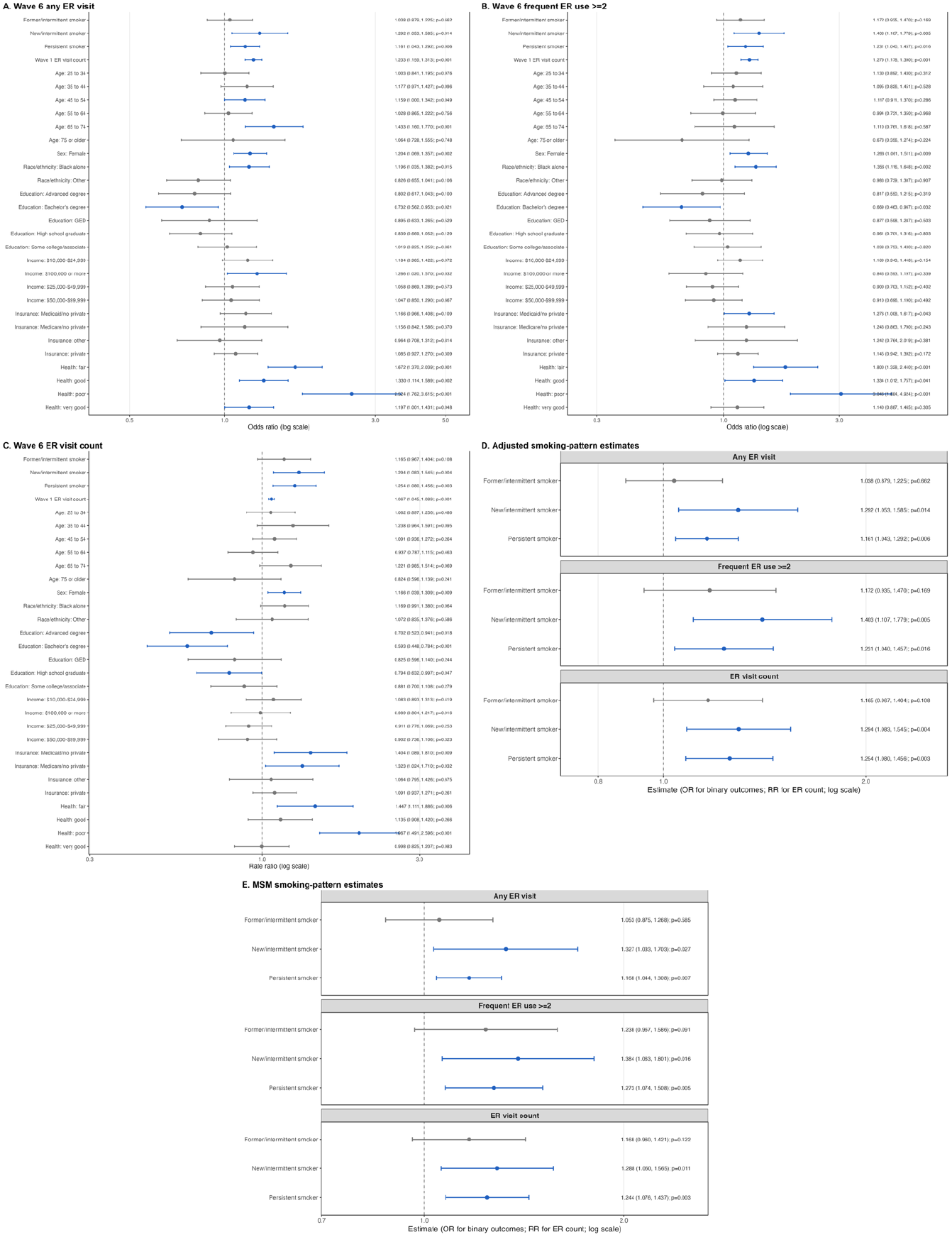
Wave 6 ER utilization and smoking-pattern predictors from waves 1-5. weighted BRR/Fay estimates were used for Wave 6 emergency room outcomes. Reference level is persistent non-smokers. Panels A-C present full adjusted models for any ER visit, frequent ER use, and ER visit count. Panels D and E summarize smoking-pattern predictors from the adjusted and marginal structural models, respectively.

Marginal structural models using stabilized inverse probability weights showed the same pattern, with significant associations among new/intermittent and persistent smokers but weaker, non-significant associations among former/intermittent smokers (Figure 3E). Together, these findings support a persistence-related association between smoking exposure and later ER utilization and highlight the potential impact of smoking cessation in reducing or eliminating the excess ER-utilization burden associated with continued smoking.

Because the former/intermittent smoking category may include both sustained quitters and individuals who relapsed, we conducted an additional cessation-focused trajectory analysis. Sustained quitters were defined as Wave 1 smokers who were non-smokers at all subsequent included waves, while relapsed/intermittent smokers were Wave 1 smokers who were not consistently non-smokers during follow-up. Sustained quitters (waves 1-5) were not significantly different from persistent non-smokers for any of the three Wave 6 ER outcomes (Supplementary Figure 3A), suggesting that sustained cessation may reduce the excess ER-utilization risk seen among continued smokers. Because Wave 6 data were collected after the onset of the COVID-19 pandemic, we also performed a sensitivity analysis using Waves 1–4 smoking patterns to predict Wave 5 ER outcomes. Results similarly showed no significant difference between sustained quitters and persistent non-smokers (Supplementary Figure 3B).

## DISCUSSION

The relationship between smoking and emergency room (ER) utilization has received relatively limited attention compared with other substance-use predictors of emergency care, and some prior analyses of ER use and substance use did not directly evaluate cigarette smoking (Capp et al., 2014; Walley et al., 2012). To our knowledge, this study is among the first nationally representative longitudinal analyses to examine smoking status, smoking intensity, and smoking trajectories in relation to subsequent ER utilization among U.S. adults. The use of PATH survey weights and replicate weights allowed us to account for the complex sampling design (Hyland et al., 2017; Opsomer et al., 2024). Our repeated-wave structure design further allowed us to evaluate temporal associations, multiple ER outcomes, and smoking trajectories over time rather than relying on a single cross-sectional smoking measure.

Our findings suggest that smoking may contribute to emergency care burden not only by increasing the likelihood of any ER use, but also by increasing recurrent and higher-volume utilization, particularly among heavier smokers. In adjusted models, current established smoking was associated with a 26.4% higher rate of subsequent ER visits, with higher rates among smokers consuming <20 cigarettes/day and *≥*20 cigarettes/day compared with non-smokers. The per-cigarette association was modest, but scaled to one pack/day, it corresponded to approximately 20% higher ER visit rates. Given the high cost of emergency care, these differences may be meaningful at the population level: national estimates suggest that ED visit costs totaled $76.3 billion across 144.8 million visits in 2017, and more recent claims-based data show continued increases in ER spending and prices (Moore & Liang, 2020; Health Care Cost Institute, 2023). These findings suggest that reducing smoking exposure may have implications beyond long-term chronic disease prevention, potentially lowering recurrent acute-care use and its associated healthcare burden.

These findings are consistent with prior evidence linking tobacco use to greater acute healthcare utilization. Among non-Hispanic Black and Hispanic men aged 40 years or older with chronic conditions, exclusive cigarette smoking was associated with higher odds of past-year ED use compared with non-tobacco use (AOR 1.66, 95% CI [1.25–2.19]), with stronger associations among dual tobacco users (AOR 1.75, 95% CI [1.23–2.50]) and poly-tobacco users (AOR 4.10, 95% CI [2.46–6.84]) (Merianos et al., 2023). Similarly, a matched cohort study found that smokers had higher rates of all-cause ED visits (RR 1.08), smoking-related ED visits (RR 1.78), all-cause hospitalizations (RR 1.31), and smoking-related hospitalizations (RR 2.23) than non-smokers (Baliunas et al., 2023). ED-based studies also show a disproportionate tobacco burden among emergency care populations: current tobacco use was reported by 26–27% of ED patients overall and by approximately 36% of patients younger than 50 years in a Canadian ED study (Smith, 2011). Smoking-attributable illness has also been estimated to account for 4.9% of adult ED visits, 6.8% of ED admissions, and 10.0% of hospital charges (Bernstein et al., 2002). Together, these studies consistently support our finding.

Our trajectory findings suggest that both persistent smoking and new/intermittent smoking are associated with higher later ER utilization, supporting the importance of repeated smoking exposure over time rather than a single baseline smoking measure. This pattern is consistent with trajectory-based studies showing that sustained smoking exposure is associated with cumulative health risk. In a Korean cohort of 60,709 young men with smoking measured seven times, very-high steady smokers had a 49% higher risk of ASCVD than low-steady smokers, with higher risks of ischemic heart disease (HR 1.63), total stroke (HR 1.36), and ischemic stroke (HR 1.58) (Jee et al., 2019). Similarly, in a population-based cohort of 2,448,548 men with smoking measured across four waves, steady heavy smokers had higher cancer incidence (HR 1.53, 95% CI [1.49–1.58]) and cancer mortality (HR 2.64, 95% CI [2.50–2.79]) than steady non-smokers (Luu et al., 2022). The elevated ER utilization among new/intermittent smokers in our study may reflect smoking initiation, re-initiation, or unstable smoking behavior, suggesting that even emerging or intermittent smoking exposure may contribute to acute-care burden. This is consistent with prior evidence linking active smoking, smoking relapse, and smoking-related disease burden with higher ED and healthcare utilization (Baliunas et al., 2023; Kao et al., 2019).

In contrast, former/intermittent smokers and sustained quitters were not significantly different from persistent non-smokers in later ER utilization, suggesting that stopping smoking may attenuate the excess risk seen among persistent and new/intermittent smokers. This interpretation is supported by evidence that cessation reduces smoking-related morbidity over time and that healthcare utilization among former smokers may move closer to never-smoker patterns than current-smoker patterns (Fishman et al., 2003; U.S. Surgeon General, 2020). Together, these findings suggest that sustained smoking exposure increases later ER burden, whereas cessation may reduce this excess burden toward the level observed among persistent non-smokers.

Our findings support integrating evidence-based tobacco cessation into routine clinical care, including ER and urgent care encounters. The USPSTF recommends that clinicians ask all adults about tobacco use, advise them to stop, and provide behavioral interventions and FDA-approved pharmacotherapy for cessation (U.S. Preventive Services Task Force, 2021). In acute-care settings, brief bedside counseling, nicotine replacement therapy initiation, and quitline referral may provide feasible opportunities to reach smokers who may otherwise have limited access to preventive care (CDC, 2024). At the policy level, comprehensive tobacco-control programs, smoke-free laws, tobacco price increases, and mass-reach health communication campaigns remain recommended strategies to reduce smoking prevalence and smoking-related morbidity (Community Preventive Services Task Force, 2024).

Despite the novel findings, the study is not without limitations. Public-use PATH data often lack granular geographic location measures. If ZIP code or census-tract information were available, future work could incorporate SES measures such as the CDC/ATSDR Social Vulnerability Index (SVI) (CDC/ATSDR, 2026) by linking with SDOH datasets. Restricted geographic data could also enable linkage to local healthcare access measures, including primary care provider density, urgent care availability, federally qualified health centers, rural health clinics, and hospital/ER proximity. These are crucial factors for emergency department use, with higher acute-care use in primary care shortage areas and lower use where clinic density or urgent care access is greater (Patel et al., 2024; Mathison et al., 2013), which helps clarify whether the association between smoking and ER usage reflects smoking behavior itself or broader social disadvantage. Additionally, although the repeated-wave design supports temporal ordering, residual confounding and reverse causation remain possible. ER use is also a nonspecific outcome: visits may reflect smoking-related illness, but may also be due to accidents, injuries, abdominal pain, acute respiratory infections, headaches, or other conditions not directly attributable to smoking (HCUP, 2018). Finally, smoking behavior and ER utilization were self-reported, which may introduce recall, perception, or reporting bias.

## CONCLUSIONS

Smoking, heavier smoking intensity, and persistent or emerging smoking patterns were associated with higher subsequent emergency room (ER) utilization across multiple outcomes. Findings support a potential causal relationship between smoking exposure and later ER use. Smoking cessation may attenuate the excess acute-care burden associated with continued or newly emerging smoking to the level similar to never-smokers. These findings indicate that smoking should be considered not only a risk factor for long-term chronic disease, but also a predictor of acute health-care utilization. Integrating cessation counseling, nicotine replacement therapy, quitline referral, and preventive care linkage into ER and urgent care settings, alongside broader tobacco-control policies, may help reduce smoking-related healthcare burden.

## SUPPLEMENTARY MATERIAL

A Contributorship Form detailing each author’s specific involvement with this content, as well as any supplementary data, are available online.

## DATA AVAILABILITY STATEMENT

The data used in this study are publicly available and can be accessed through the Population Assessment of Tobacco and Health (PATH) Study website. Details on how to access the data are available at https://pathstudyinfo.nih.gov/. Researchers can request access following the repository’s guidelines.

## SUPPLEMENTARY FIGURES

**Supplementary Figure 1.**
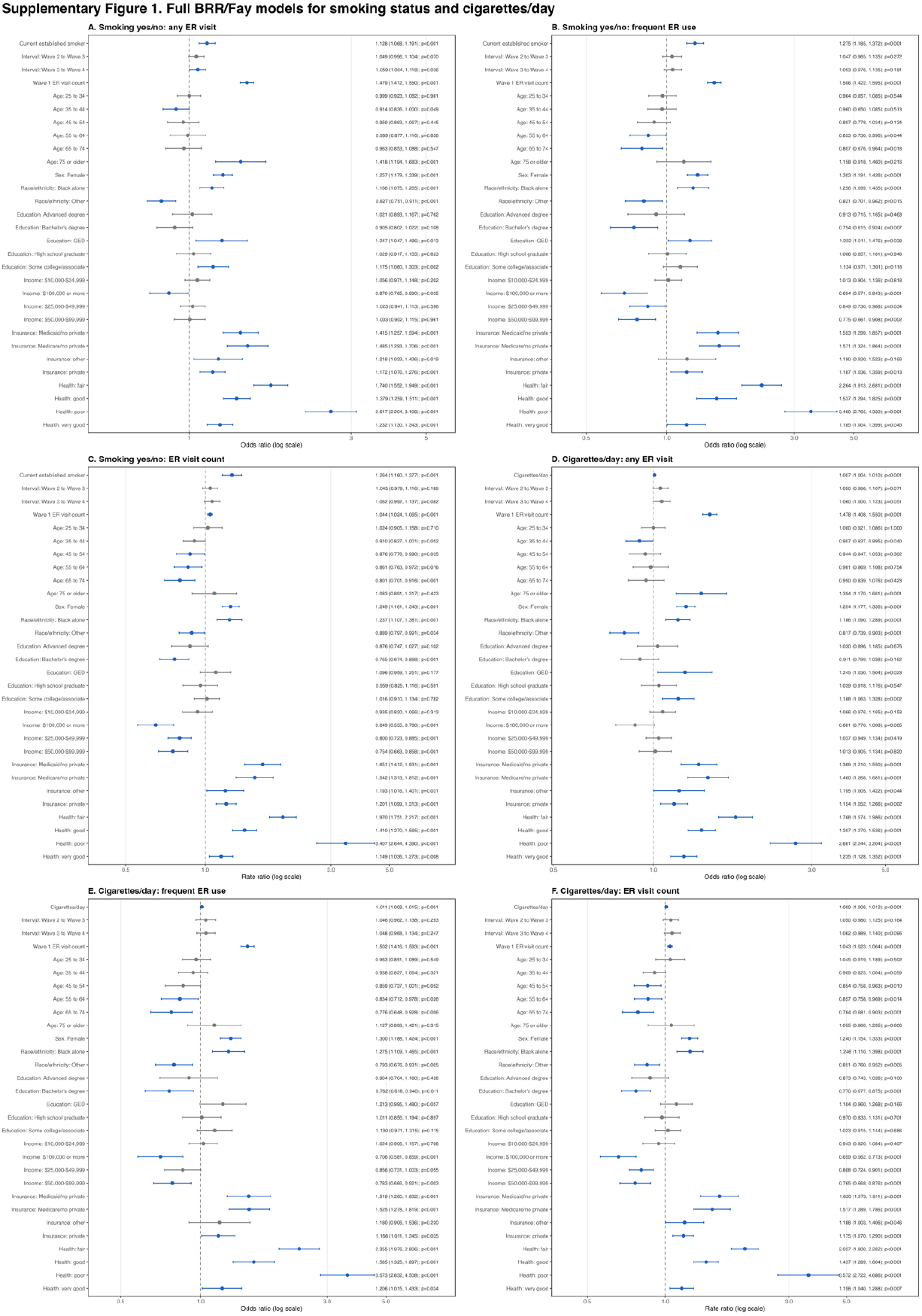
Full adjusted models for smoking status, cigarettes/day, and next-wave ER utilization. shows full adjusted weighted BRR/Fay regression models for smoking yes/no and cigarettes/day predictors across three next-wave ER outcomes. Panels A-C show models using current established smoking status, and Panels D-F show models using cigarettes/day. Models adjust for interval, Wave 1 ER visit count, age, sex, race/ethnicity, education, income, insurance, and self-rated physical health. Points represent adjusted estimates, horizontal bars represent 95% confidence intervals, and dashed vertical lines indicate the null value of 1.0.

**Supplementary Figure 2.**
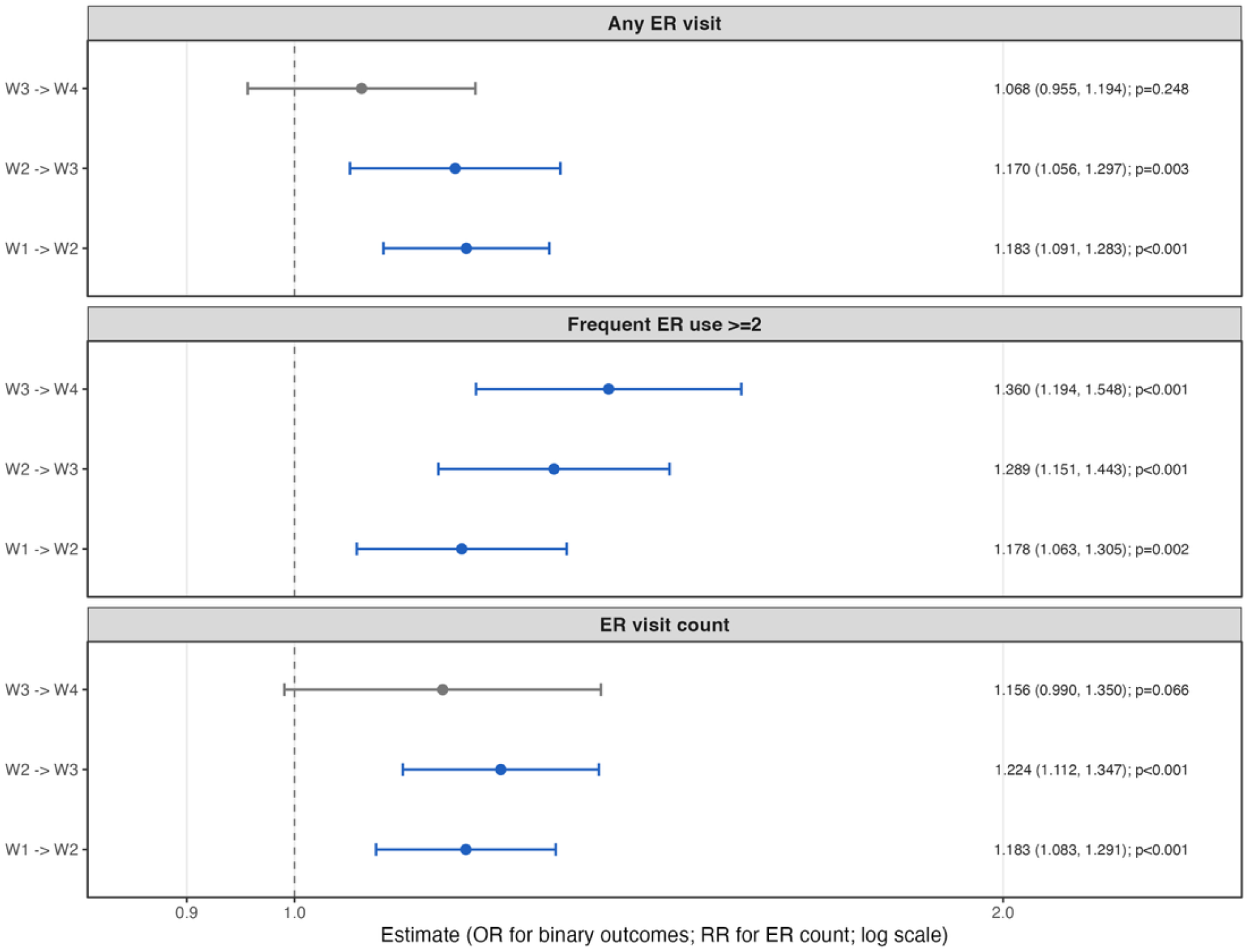
Propensity score-matched estimates for smoking and next-wave emergency room utilization. Supplementary Figure 2 shows BRR/Fay propensity score-matched estimates for smoking and subsequent emergency room (ER) outcomes across survey-wave intervals. Estimates are shown for any ER visit, frequent ER use, and ER visit count. Points represent effect estimates, horizontal bars represent 95% confidence intervals, and dashed vertical lines indicate the null value of 1.0.

**Supplementary Figure 3.**
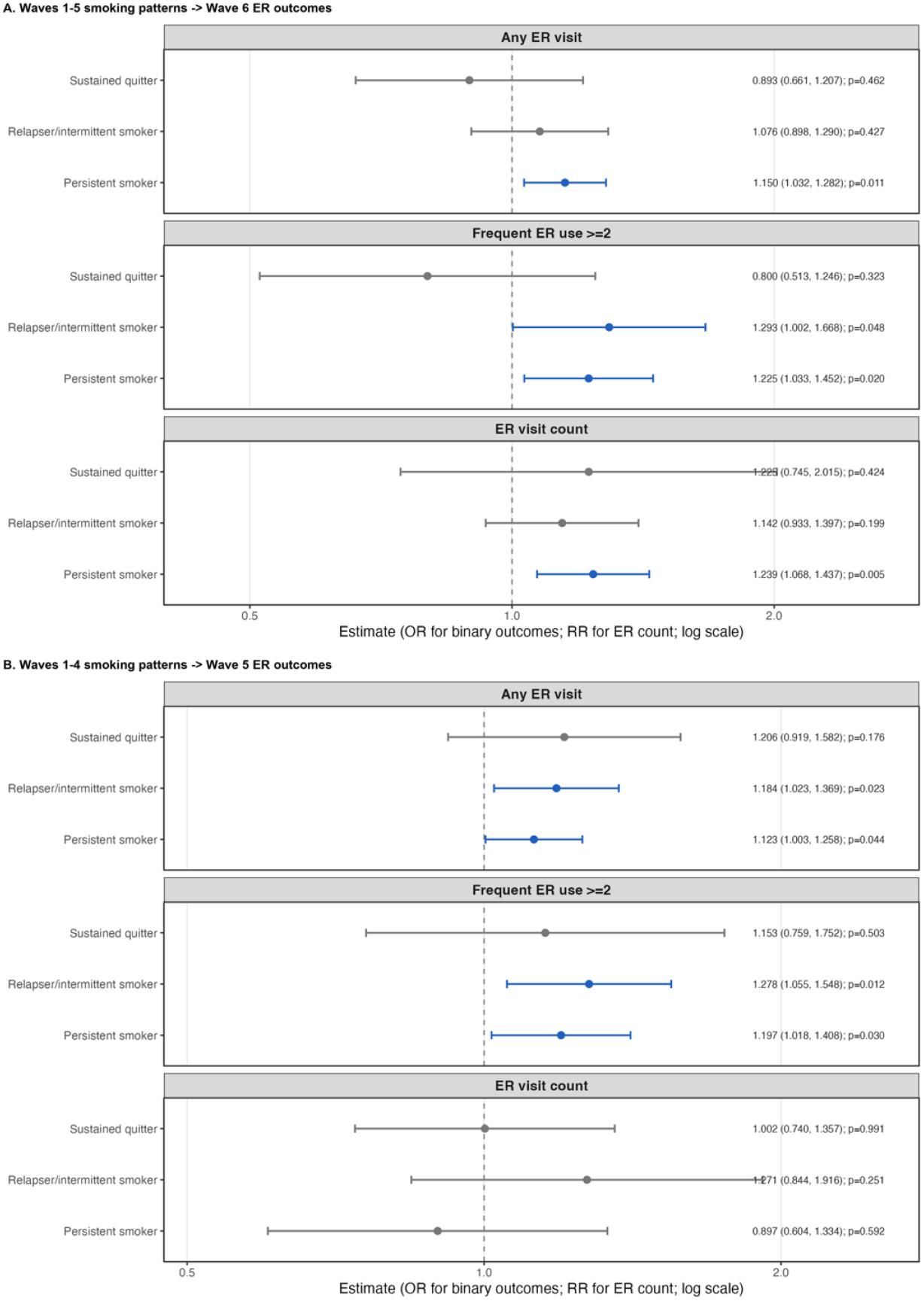
ER outcomes by sustained quitter/ relapse/intermittent smoking pattern. shows weighted BRR/Fay estimates for associations between sustained quitting/smoking-pattern groups and ER outcomes measured at Wave 6 and Wave 5. Panel A presents estimates for Wave 6 ER outcomes (predictors waves1-5), and Panel B presents estimates for Wave 5 ER outcomes (predictors waves 1-4). Persistent non-smokers are the reference group. Points represent adjusted estimates, horizontal bars represent 95% confidence intervals, and dashed vertical lines indicate the null value of 1.0.

## REFERENCES

1. Agency for Healthcare Research and Quality. (2021). Most frequent reasons for emergency department visits, 2018. HCUP Statistical Brief No. 286. https://hcup-us.ahrq.gov/reports/statbriefs/sb286-ED-Frequent-Conditions-2018.jsp

2. American Cancer Society. (2024). Benefits of quitting smoking over time. https://www.cancer.org/cancer/risk-prevention/tobacco/guide-quitting-smoking/benefits-of-quitting-smoking-over-time.html

3. Arrazola, R. A., et al. (2025). Tobacco product use among adults: United States, 2017–2023. Morbidity and Mortality Weekly Report. https://www.cdc.gov/mmwr/volumes/74/wr/mm7407a3.htm

4. Baliunas, D., Selby, P., de Oliveira, C., Kurdyak, P., Rosella, L., Zawertailo, L., Fu, L., & Sutradhar, R. (2023). Primary care-based smoking cessation treatment and subsequent healthcare service utilisation: A matched cohort study of smokers using linked administrative healthcare data. Tobacco Control, 32(1), 72–79. 10.1136/tobaccocontrol-2021-056522

5. Barmparessou, Z., et al. (2020). Smoking habit among adults visiting the emergency room for an asthma exacerbation. Tobacco Prevention & Cessation, 6(Supplement). 10.18332/tpc/128487

6. Beard, E., Shahab, L., Cummings, K. M., Michie, S., & West, R. (2013). Short-term changes in health and well-being after smoking cessation: A prospective cohort study. BMJ Open, 3(11), e003310. 10.1136/bmjopen-2013-003310

7. Bernstein, S. L. (2002). Smoking-related illness in the emergency department: An attributable risk model. American Journal of Emergency Medicine, 20(3), 161–164. https://pubmed.ncbi.nlm.nih.gov/11992333/

8. Bernstein, S. L. (2002). Preventive care in the emergency department: Diagnosis and management of smoking and smoking-related illness in the emergency department. https://pubmed.ncbi.nlm.nih.gov/12093715/

9. Bernstein, S. L., et al. (2011). A randomized trial of a multicomponent cessation strategy for emergency department smokers. https://pubmed.ncbi.nlm.nih.gov/21676054/

10. Cairns, C., Ashman, J. J., & Kang, K. (2024). Emergency department visit rates by selected characteristics: United States, 2022. NCHS Data Brief, 503, 1–8. https://www.cdc.gov/nchs/products/databriefs/db503.htm

11. Campagna, D., et al. (2024). Tobacco smoking or nicotine phenotype and severity of clinical presentation at the emergency department (SMOPHED): Protocol for a noninterventional observational study. JMIR Research Protocols, 13, e54041. 10.2196/54041

12. Capp, R., Kelley, L., Ellis, P., Carmona, J., Lofton, A., Cobbs-Lomax, D., & D’Onofrio, G. (2016). Reasons for frequent emergency department use by Medicaid enrollees: A qualitative study. Academic Emergency Medicine, 23(4), 476–481. 10.1111/acem.12952

13. Castner, J., Wu, Y. W. B., Mehrok, N., Gadre, A., & Hewner, S. (2015). Frequent emergency department utilization and behavioral health diagnoses. Nursing Research, 64(1), 3–12. 10.1097/NNR.0000000000000065

14. Centers for Disease Control and Prevention. (2024a). Burden of cigarette use in the U.S. https://www.cdc.gov/tobacco/campaign/tips/resources/data/cigarette-smoking-in-united-states.html

15. Centers for Disease Control and Prevention. (2024b). Health and economic benefits of tobacco use interventions. https://www.cdc.gov/tobacco/php/data-statistics/economic-trends/index.html

16. Centers for Disease Control and Prevention. (2024c). Benefits of quitting smoking. https://www.cdc.gov/tobacco/about/benefits-of-quitting.html

17. Centers for Disease Control and Prevention. (2024d). Clinical interventions for tobacco cessation. https://www.cdc.gov/tobacco/hcp/patient-care-settings/clinical.html

18. Centers for Disease Control and Prevention/Agency for Toxic Substances and Disease Registry. (2026). CDC/ATSDR Social Vulnerability Index. https://www.atsdr.cdc.gov/place-health/php/svi/index.html

19. Centers for Disease Control and Prevention, National Center for Health Statistics. (2024). Fast-Stats: Emergency department visits. https://www.cdc.gov/nchs/fastats/emergency-department.htm

20. Cheung, P. T., Wiler, J. L., Lowe, R. A., & Ginde, A. A. (2012). National study of barriers to timely primary care and emergency department utilization among Medicaid beneficiaries. Annals of Emergency Medicine, 60(1), 4–10.e2. https://pubmed.ncbi.nlm.nih.gov/22418570/

21. Community Preventive Services Task Force. (2024). Tobacco use and secondhand smoke exposure. https://www.thecommunityguide.org/topics/tobacco.html

22. Fishman, P. A., Khan, Z. M., Thompson, E. E., & Curry, S. J. (2003). Health care costs among smokers, former smokers, and never smokers in an HMO. Health Services Research, 38(2), 733– 749. https://pubmed.ncbi.nlm.nih.gov/12785570/

23. Health Care Cost Institute. (2023). Emergency room spending, price, and use trends 2012–2021. https://healthcostinstitute.org/all-hcci-reports/emergency-room-spending-price-and-use-trends-2012-2021/

24. Hyland, A., Ambrose, B. K., Conway, K. P., et al. (2017). Design and methods of the Population Assessment of Tobacco and Health (PATH) Study. Tobacco Control, 26(4), 371–378. 10.1136/tobaccocontrol-2016-052934

25. Jee, Y., Jeon, J., Back, J. H., Ryu, M., & Cho, S. (2019). Trajectory of smoking and incidence of atherosclerotic cardiovascular disease among Korean young adult men. International Journal of Environmental Research and Public Health, 16(12), 2219. 10.3390/ijerph16122219

26. Kao, W. H., et al. (2019). Smoking relapse and type 2 diabetes mellitus-related emergency department visits among senior patients with diabetes. Preventing Chronic Disease, 16, E27. https://www.cdc.gov/pcd/issues/2019/19_0027.htm

27. Lanari, D., Pasqualini, M., & Pieroni, L. (2022). Is it time to quit? Smoking persistence and selfrated health. Journal of Population Ageing, 15(2), 363–380. 10.1007/s12062-020-09292-5

28. Liang, K. Y., & Zeger, S. L. (1986). Longitudinal data analysis using generalized linear models. Biometrika, 73(1), 13–22. 10.1093/biomet/73.1.13

29. Luu, X. Q., Han, M. A., Bui, T. K., Tran, T. T., Lim, M. K., & Oh, J.-K. (2022). Smoking trajectory and cancer risk: A population-based cohort study. Tobacco Induced Diseases, 20, 1–10. 10.18332/tid/152137

30. Madani, N. A., & Carpenter, D. O. (2023). Patterns of emergency room visits for respiratory diseases in New York State in relation to air pollution, poverty and smoking. International Journal of Environmental Research and Public Health, 20(4), 3267. 10.3390/ijerph20043267

31. Mathison, D. J., et al. (2013). Primary care spatial density and nonurgent emergency department utilization. Pediatrics, 131(6), e1751–e1759. 10.1542/peds.2012-2345

32. Merianos, A. L., et al. (2023). Current tobacco use patterns associated with healthcare utilization among non-Hispanic Black and Hispanic men with chronic conditions. Addictive Behaviors, 143, 107695. https://pubmed.ncbi.nlm.nih.gov/37001260/

33. Moore, B. J., & Liang, L. (2020). Costs of emergency department visits in the United States, 2017. HCUP Statistical Brief No. 268. Agency for Healthcare Research and Quality. https://hcup-us.ahrq.gov/reports/statbriefs/sb268-ED-Costs-2017.jsp

34. Mullen, K. A., et al. (2017). Effectiveness of a hospital-initiated smoking cessation programme: 2-year health and healthcare outcomes. Tobacco Control. https://pubmed.ncbi.nlm.nih.gov/27225016/

35. National Center for Health Statistics. (2026). Cigarette and electronic cigarette use among adults by urbanization level: United States, 2024. https://www.cdc.gov/nchs/data/hestat/hestat115.htm

36. National Institute on Drug Abuse. (n.d.). Population Assessment of Tobacco and Health (PATH) Study data access. https://nida.nih.gov/research/nida-research-programs-activities/population-assessment-tobacco-health-path-study/data-access

37. Opsomer, J. D., et al. (2024). Update to the design and methods of the Population Assessment of Tobacco and Health (PATH) Study. Tobacco Control, 33(6), 733–738. https://pubmed.ncbi.nlm.nih.gov/37258274/

38. Patel, S. Y., Baum, A., & Basu, S. (2024). Geographic variations and facility determinants of acute care utilization and spending for ambulatory care-sensitive conditions. American Journal of Managed Care, 30(11), e329–e336. 10.37765/ajmc.2024.89630

39. Pettit, N., et al. (2022). Tobacco cessation interventions in emergency care settings. https://link.springer.com/article/10.1186/s44201-022-00006-5

40. Pope, I., et al. (2024). Cessation of Smoking Trial in the Emergency Department (COSTED): A multicentre randomised controlled trial. Emergency Medicine Journal, 41(5), 276–282. 10.1136/emermed-2023-213824

41. Robins, J. M., Hernán, M. A., & Brumback, B. (2000). Marginal structural models and causal inference in epidemiology. Epidemiology, 11(5), 550–560. https://pubmed.ncbi.nlm.nih.gov/10955408/

42. Rosenbaum, P. R., & Rubin, D. B. (1983). The central role of the propensity score in observational studies for causal effects. Biometrika, 70(1), 41–55. 10.1093/biomet/70.1.41

43. Selya, A., et al. (2020). Smoking is associated with a higher risk of unplanned medical visits among adult patients with diabetes. BMC Health Services Research, 20, 415. 10.1186/s12913-020-05277-4

44. Smith, P. M. (2011). Tobacco use among emergency department patients. International Journal of Environmental Research and Public Health, 8(1), 253–263. 10.3390/ijerph8010253

45. Udalova, V., Powers, D., Robinson, S., & Notter, I. (2022). Most vulnerable more likely to depend on emergency rooms for preventable care. U.S. Census Bureau. https://www.census.gov/library/stories/2022/01/who-makes-more-preventable-visits-to-emergency-rooms.html

46. U.S. Department of Health and Human Services. (2020). Smoking cessation: A report of the Surgeon General. https://www.hhs.gov/surgeongeneral/reports-and-publications/tobacco/2020-cessation-sgr-factsheet-key-findings/index.html

47. U.S. Preventive Services Task Force. (2021). Interventions for tobacco smoking cessation in adults, including pregnant persons: U.S. Preventive Services Task Force recommendation statement. JAMA, 325(3), 265–279. 10.1001/jama.2020.25019

48. VanFrank, B., et al. (2024). Adult smoking cessation: United States, 2022. Morbidity and Mortality Weekly Report. https://www.cdc.gov/mmwr/volumes/73/wr/mm7329a1.htm

49. Walley, A. Y., Paasche-Orlow, M., Lee, E. C., Forsythe, S., Chetty, V. K., Mitchell, S., & Jack, B. W. (2012). Acute care hospital utilization among medical inpatients discharged with a substance use disorder diagnosis. Journal of Addiction Medicine, 6(1), 50–56. 10.1097/ADM.0b013e318231de51

50. World Health Organization. (2026). Tobacco and nicotine fact sheet. https://www.who.int/news-room/fact-sheets/detail/tobacco

51. Yale Medicine. (n.d.). Tobacco cessation in the emergency department. https://www.yalemedicine.org/conditions/tobacco-cessation-in-the-emergency-department

52. Zhou, R. A., Baicker, K., Taubman, S., & Finkelstein, A. N. (2017). The uninsured do not use the emergency department more: They use other care less. Health Affairs, 36(12), 2115–2122. 10.1377/hlthaff.2017.0218

